# Leveraging AutoML to provide NAFLD screening diagnosis: Proposed machine learning models

**DOI:** 10.1101/2020.10.20.20216291

**Authors:** Ali Haider Bangash

**Affiliations:** Shifa College of Medicine, Shifa Tameer e Millat University, Islamabad, Pakistan

**Keywords:** NAFLD, Fatty liver disease, autoML, machine learning, Extreme Gradient Boosting, Xgboost, LightGBM, Random Forest, Regularized Greedy Forest, Extra Trees, k-Nearest Neighbors, KNNs, Logistic Regression, Neural Network, Ensembling, COVID 19

## Abstract

NAFLD is reported to be the only hepatic ailment increasing in its prevalence concurrently with both; obesity & T2DM. In the wake of a massive strain on global health resources due to COVID 19 pandemic, NAFLD is bound to be neglected & shelved. Abdominal ultrasonography is done for NAFLD screening diagnosis which has a high monetary cost associated with it. We utilized MLjar, an autoML web platform, to propose machine learning models that require no coding whatsoever & take in only easy-to-measure anthropometric measures for coming up with a screening diagnosis for NAFLD with considerably high AUC. Further studies are suggested to validate the generalization of the presented models.

## 1 Introduction

Hepatic diseases taking the lives of as many as 1.75 million people worldwide annually are a menace to be reckoned with.^(1)^ Younossi ZM et al^(1)^ indicated that non-alcoholic fatty liver disease (NAFLD) is the only hepatic pathology growing exponentially in its prevalence, in coincidence with the increasing rates of both; obesity as well as type 2 diabetes mellitus (T2DM): all wreaking havoc.^(2)(3)(4)^ Where COVID-19 pandemic is undeniably encroaching over the global health system’s resources, NAFLD, along with other ailments, is bound to be ignored which may lead to a rise in the associated morbidity & mortality.

A considerable amount of financial resources are being put in globally in the fight against COVID-19 pandemic where radiological diagnosis for NAFLD demands a considerable allocation of funds, the prioritization of resources shall undeniably end up in missing cases on the cost of the patients as well as their families in terms of the psychological burden. A low-cost, accurate system to screen patients for a potential NAFLD diagnosis using simple, easy-to-measure anthropometric measures is a dire need of time, thus.

Huang BX et al, in a cross-sectional study conducted in the Health Examination Center in Guangzhou, China took in anthropometric measures & abdominal ultrasonography & concluded neck circumference to be an independent predictor for the fatty liver disease.^(5)^

The purpose of this study is to create such machine learning models that take in the said easy-to-measure variables from the Huang BX et al study^(5)^ & come up with an autoML protocol for initial screening diagnosis for NAFLD: models that are, if not having a potential to replace, should be at least comparable with the abdominal ultrasound screening diagnosis for NAFLD. By adopting Mljar^(6)^, a zero-code autoML web platform providing feature preprocessing and eningeering, algorithm training and hyperparameters selection bundle for machine learning, we are able to create such practical models.

## 2 Materials & Methods

### 2.1 Study Cohort

The study takes in data^(7)^ from Huang BX et al.^(5)^. The authors took in 4053 subjects, 2436 men and 1617 women between 20 and 88 years of age, after excluding those patients that had a history of co-morbid conditions as well as those with a lack of heaptic ultrasonography data. Patients’ history records were inquired & state-of-the-art methods were adopted to measure anthropometric & biochemical variables leaving negligible measurement errors, only. Contrary to South Asian standards where BMI of ≥ 25 Kg/m^2^ is termed as Overweight, a BMI ≧24 kg/m^2^, for both genders, is termed as Overweight by Huang BX et al.^(5)^, citing Zhou B^(8)^. The Graif’s criteria^(9)^ was adopted to diagnose Fatty liver disease on ultrasonography.

### 2.2 Statistical analysis

#### Development of Models

##### Homogenous Development Framework

MLjar^(6)^, an AutoML zero-code machine learning web platform, is a complete package for data loading, pre-processing, modelling & result interpretation with a considerably high quality of machine learning models which can be deployed both locally & across a rest API.

A homogenous approach (Table 1) was adopted for the development of the models vis-à-vis the preprocessing & tuning protocols as well as system specifications so as to keep the model development bias to a minimum.

**Table 1.** Homogenous Development Framework

| Development framework | Features |
| --- | --- |
| Preprocessing protocol | One-hot encoding to convert categorical features |
| System specifications | 8 CPUs, 15 GB RAM |
| Tuning protocol<br>Validation type<br>Tuning mode<br>Time limit for single model | 15-fold cross-validation, Shuffling of samples & stratification of classes in folds<br>Perfect mode (25-35 models)<br>5 minutes |

##### Models developed

Henceforth mentioned machine learning models were, thus, created:

- Extreme Gradient Boosting (Xgboost)
- LightGBM (LGBM)
- Random Forest (RF)
- Regularized Greedy Forest (RGF)
- Extra Trees (ET)
- k-Nearest Neighbor (KNN)
- Logistic Regression (LR)
- Neural Network (NN)

#### Outcome variables

Since the class imbalance was considerably high, the discriminative ability of the models were the primary outcome variables. AUC-ROC analysis was adopted to measure that ability.^(10)^ The respective values were interpreted in accordance with the schema provided by Lau L et al.^(10)^ (Table 2)

**Table 2.** Interpretation of the AUC-ROC analysis (Adopted from: Lau L et al.) (AUC-ROC: Area under the receiver operating characteristic curve)

| AUC-ROC value | Interpretation |
| --- | --- |
| >0.9 | Excellent discrimination |
| >0.75 | Good discrimination |
| >0.5 | Random guessing |

Secondarily, training time was also analyzed. Ideally, the best model shall be the one that has the highest discriminating capacity & yields results within the smallest time period.

## 3 Results

### 3.1 Algorithm performances

The study adopted a zero-code ML platform to come up with 8 types of machine learning models that take in easy-to-measure anthropometric measures such as BMI & waist-to-hip ratio in order to provide a screening diagnosis for NAFLD.

As indicated in table 3, all of the algorithms, trained in accordance with the aforementioned Homogenous Development Framework, have good discriminating ability to designate the dichotomous variable of interest.

**Table 3.** AUC & training time of the proposed models

| Algorithm | Area Under Curve | Time | Ensemble Averaging | Time |
| --- | --- | --- | --- | --- |
| Xgboost | 0.822004 | 0:07:45 | 0.822679 | 0:00:35 |
| LGBM | 0.823532 | **0:10:05** | - | - |
| RF | **0.824809** | 0:04:09 | 0.825137 | 0:00:33 |
| RGF | 0.823396 | 0:00:10 | **0.826389** | 0:00:35 |
| ET | 0.823557 | 0:01:02 | 0.824575 | 0:00:35 |
| KNN | **0.815769** | 0:00:06 | 0.816616 | 0:00:28 |
| LR | 0.821363 | **0:00:01** | 0.821632 | 0:00:26 |
| NN | 0.820271 | 0:03:16 | - | - |

RF came out to have the highest discriminating ability, with a computation time of 4 minutes 9 seconds. Out of the proposed models, KNN had the least AUC but a considerably less computation time of only 6 seconds. LGBM required as much as 10 minutes to come up with a considerable AUC. LR completed its computation in the least amount of time.

Among the ensemble averages, RGF achieved the highest average. Given that the best model of RGF was up with its training in only 10 seconds & an additional 35 seconds were lapsed for ensemble averaging, RGF outperforms all others models by achieving the highest AUC & thus exhibiting the best discriminating ability out of all the models.

KNN exhibited the lowest ensemble average. (Ensemble average of LGBM & NN was not calculated.) Thus, it is safe to indicate that KNN underperformed the most out all the proposed models.

## 4 Discussion

Many studies have been done to utilize machine learning for the prediction of fatty liver disease. Atabaki-Pasdar N et al^(11)^ in a major modelling & validation study concluded that the highest AUC (of 0.84 for the respective study) is obtained by the combination of “-omics” data & clinical variables. Using MRI-derived proton density fat fraction for referencing, Han A et al^(12)^ developed deep learning one-dimensional convolutional neural networks for NAFLD diagnosis by taking in ultrasound data.^1^ By taking in all the patients who had been screened for fatty liver at the New Taipei City Hospital between the 1st and 31^st^ of December 2009, Wu CC et al^(13)^ developed several classification models to predict fatty liver disease and obtained the highest AUC of 0.925 on a Random Forest model. Feature selection was employed to obtain the best variables to be fed in the models, here. The utilization of machine learning to predict hepatic pathologies in general & NAFLD in particular is thus evident.

Our proposed models are the very first effort, to the best of our knowledge, to leverage autoML zero-code platforms to come up with machine learning models that are trained to have a good discriminating ability to predict NAFLD using only anthropometric measures. The proposed models neither require costly analysis so that variables, such as unltrasonographic signals, may be fed in them to obtain a prediction nor does it require considerably high computation time & resources.

This been stated, the model does require external validation using data from populations different from its training population. Only thus can a machine learning model’s generalization can be truly validated. Moreover, a study comparing the presented model’s diagnosis with an abdominal ultrasound diagnosis for NAFLD, the predictions assessed against hepatic biopsy, is proposed to be in order to explore the presented models’ potential to replace abdominal ultrasound as an initial diagnostic tool for NAFLD.

Since autoML platform was adopted, the proposed models are analogous to a black-box, the internal workings of which are difficult to decipher. Moreover, the computation time of the best LR model is only 1 second which might possibly be due to overfitting of the respective model.

The presented models indicate that the fusion of machine learning & medicine is fruitful for cutting down the associated costs of screening and initial diagnosis of NAFLD: an ailment that has considerable morbidity and mortality associated with it.

## 5 Conclusion

By adopting an autoML zero-code platform, machine learning models with good discriminating ability are presented that require only easy-to-measure anthropometric measures as input variables to come up with an initial screening diagnosis for NAFLD. Further studies should be conducted to compare the proposed models with abdominal ultrasound for the screening diagnosis of NAFLD.

## Data Availability

The data used for model development has been very kindly been shared by Huang BX et al(1) at figshare. (2)
(1) Huang B, Zhu M, Wu T, Zhou J, Liu Y, Chen X, et al. Neck Circumference, along with Other Anthropometric Indices, Has an Independent and Additional Contribution in Predicting Fatty Liver Disease. Targher G, editor. PLoS One [Internet]. 2015 Feb 13 [cited 2020 Jul 13];10(2):e0118071. Available from: https://dx.plos.org/10.1371/journal.pone.0118071
(2) Neck Circumference, along with Other Anthropometric Indices, Has an Independent and Additional Contribution in Predicting Fatty Liver Disease [Internet]. [cited 2020 Sep 28]. Available from: https://figshare.com/collections/Age_adjusted_ORs_of_FLD_according_to_joint_classification_of_NC_and_other_anthropometric_measures_/2042852

## 8 Study Association

This study is neither associated with any thesis or dissertation work nor with any conference.

## Footnotes

1 For the Han A et al^(12)^ study, the metrics against which the respective proposed model was evaluated did not include AUC.

## References

1. Younossi ZM, Stepanova M, Younossi Y, Golabi P, Mishra A, Rafiq N, et al. Epidemiology of chronic liver diseases in the USA in the past three decades. Gut [Internet]. 2020 Mar 1 [cited 2020 Jul 13];69(3):564–8. Available from: https://gut.bmj.com/content/69/3/564

2. Berry EM. The Obesity Pandemic—Whose Responsibility? No Blame, No Shame, Not More of the Same. Front Nutr [Internet]. 2020 Jan 31 [cited 2020 Jul 13];7(2):2. Available from: https://www.frontiersin.org/article/10.3389/fnut.2020.00002/full

3. Ghanemi A, Yoshioka M, St-Amand J. Will an obesity pandemic replace the coronavirus disease-2019 (COVID-19) pandemic?Med Hypotheses [Internet]. 2020 Jun [cited 2020 Jul 13];144:110042. Available from: /pmc/articles/PMC7316052/?report=abstract

4. Ghosal S, Arora B, Dutta K, Ghosh A, Sinha B, Misra A. Increase in the risk of type 2 diabetes during lockdown for the COVID19 pandemic in India: A cohort analysis. Diabetes Metab Syndr Clin Res Rev. 2020 Sep 1;14(5):949–52.

5. Huang B, Zhu M, Wu T, Zhou J, Liu Y, Chen X, et al. Neck Circumference, along with Other Anthropometric Indices, Has an Independent and Additional Contribution in Predicting Fatty Liver Disease. Targher G, editor. PLoS One [Internet]. 2015 Feb 13 [cited 2020 Jul 13];10(2):e0118071. Available from: https://dx.plos.org/10.1371/journal.pone.0118071

6. MLJAR | Machine Learning Made Simple [Internet]. [cited 2020 Oct 20]. Available from: https://mljar.com/

7. Neck Circumference, along with Other Anthropometric Indices, Has an Independent and Additional Contribution in Predicting Fatty Liver Disease [Internet]. [cited 2020 Sep 28]. Available from: https://figshare.com/collections/Age_adjusted_ORs_of_FLD_according_to_joint_classification_of_NC_and_other_anthropometric_measures_/2042852

8. Zhou B. Predictive values of body mass index and waist circumference to risk factors of related diseases in Chinese adult population. Zhonghua Liu Xing Bing Xue Za Zhi [Internet]. 2002 Feb 1 [cited 2020 Oct 20];23(1):5–10. Available from: https://europepmc.org/article/med/12015100

9. Graif M, Yanuka M, Baraz M, Blank A, Moshkovitz M, Kessler A, et al. Quantitative estimation of attenuation in ultrasound video images: Correlation with histology in diffuse liver disease. Invest Radiol [Internet]. 2000 May [cited 2020 Oct 20];35(5):319–24. Available from: https://pubmed.ncbi.nlm.nih.gov/10803673/

10. Lau L, Kankanige Y, Rubinstein B, Jones R, Christophi C, Muralidharan V, et al. Machine-Learning Algorithms Predict Graft Failure After Liver Transplantation. Transplantation [Internet]. 2017 Apr 1 [cited 2020 Oct 20];101(4):e125–32. Available from: http://journals.lww.com/00007890-201704000-00025

11. Atabaki-Pasdar Id N, Ohlsson Id M, Viñuela Id A, Frau F, Pomares-Millanid H, Haidid M, et al. Predicting and elucidating the etiology of fatty liver disease: A machine learning modeling and validation study in the IMI DIRECT cohorts PLOS MEDICINE. Jagadish VangipurapuID [Internet]. [cited 2020 Jul 13];25:40. Available from: https://doi.org/10.1371/journal.pmed.1003149

12. Han A, Byra M, Heba E, Andre MP, Erdman JW, Loomba R, et al. Noninvasive diagnosis of nonalcoholic fatty liver disease and quantification of liver fat with radiofrequency ultrasound data using one-dimensional convolutional neural networks. Radiology [Internet]. 2020 May 1 [cited 2020 Jul 13];295(2):342–50. Available from: https://pubs.rsna.org/doi/abs/10.1148/radiol.2020191160

13. Wu CC, Yeh WC, Hsu WD, Islam MM, Nguyen PA (Alex), Poly TN, et al. Prediction of fatty liver disease using machine learning algorithms. Comput Methods Programs Biomed. 2019 Mar 1;170:23–9.

